# RISK OF THROMBOEMBOLISM AFTER COVID-19 VACCINATION AND COVID-19 INFECTION

**DOI:** 10.1101/2024.02.12.24302535

**Authors:** Huong Tran, Malcolm Risk, Yung-Chun Lee, Girish Nair.B, Lili Zhao

## Abstract

**Background:** Vaccine safety monitoring systems worldwide have reported cases of venous thromboembolism and arterial thromboembolism following a COVID-19 vaccination. However, evidence shows that the association between thromboembolism and SARS-CoV-2 infection is stronger, compared to SARS-CoV-2 vaccination. Hence, weighing the risks and benefits of vaccination should also encounter the roles of vaccination in reducing infection rate, and potentially indirectly lowering the risk of thromboembolism caused by infection.

**Methods:** We conducted a self-controlled case series study (SCCS) from Dec 1^st^ 2020 to 31^st^ August 2022 (before the bivalent vaccine was available) to examinate the association between the first two doses Pfizer/Moderna vaccination and thrombotic events among patients in Corewell Health East (CHE, formerly known as Beaumont Health) healthcare system. We also investigated the effect SARS-CoV-2 infection on the risk of thrombosis events and observed a significant increased risk using the SCCS design. However, because of misclassification bias, SCCS indeed overestimated incidence rate ratio (IRR) of acute event after infection, we then proposed a case-control study addressing this misclassification issues and obtained odd ratio comparing effect of exposure on thrombosis and a subset of controls group. Finally, we analyzed the risk of thromboembolism between vaccinated and unvaccinated groups by a simple diagram, explaining possible factors that affects the probability of experiencing an acute thromboembolism event after a COVID-19 vaccination.

**Results:** Using EHR data at Corewell East, we found an increased risk of thrombosis after the first two doses of COVID-19 vaccination, with incidence rate ratios after the first dose is 1.16 (CI: [1.04, 1.29]), and after the second dose of 1.19 (CI: [1.07,1.32]). The association between thromboembolism and SARS-Cov-2 infection depends on prior vaccination status, as the conditional OR among unvaccinated and vaccinated groups are 1.77 (CI: [1.48, 2.1]) and 1.34 (CI: [1.09, 1.66]) respectively. Encountering the vaccine efficacy (VE), receiving the COVID-19 vaccine decreases the risk of thromboembolism, and the benefits of COVID-19 vaccines are much stronger in the period of high infection rate.

## 1. Introduction

COVID-19 disease has unfolded as an unprecedented global health crisis, which has significantly high transmissibility leading to the rapid spread of the virus within communities (1). The recent battle against COVID-19 pandemic has led to a race to develop effective vaccines, which have played a vital role in curbing the spread the virus and preventing severe illness. There are 81.4% (data as of 11/17/2023 – CDC (2)) of US population has been vaccinated by at least one dose and 69.5% has completed primary series for COVID-19 vaccination. While the benefits of vaccination are widely acknowledged, concerns have emerged regarding the development of acute thrombosis after vaccination (3). More specifically, some cases of venous thromboembolism followed by a mRNA-based vaccination has been reported in 2021 (4), (5), (6), (7), drawing an attention to potential risk of thrombosis after the first dose of vaccination. Some studies have confirmed that there is an increased risk of thromboembolism, ischemic stroke and cerebral venous sinus thrombosis after the first dose of BNT162b2 (Pfizer) (8), while another retrospective cohort study using EHR data also confirmed an increased risk of cerebral venous thrombosis and portal vein thrombosis after a mRNA vaccination (9). Moreover, a recent systematic review (10) has shown that the most frequent adverse event followed by a mRNA vaccination is thromboembolism. Despite those findings, vaccination is still recommended to reduce the likelihood of COVID-19 infection and its severity, (8), (11). Furthermore, it has been proven that risk of thromboembolism is strongly associated with COVID-19 (12), (13), (14), implying a higher risk of mortality (15). Therefore, study the association between thromboembolism acute events and SARS-Cov-2 vaccination should incorporate infection.

In the context of vaccine safety, the self-controlled case series (SCCS) method is particularly valuable when evaluating the causal relationship between vaccines and acute adverse events. In the 28 days following a mRNA-based vaccine, SCCS conducted using various data sources from Scotland (16), England (8), France (17), Nordic countries (18), and Hongkong (19) has consistently confirmed the positive correlation between thromboembolism and a mRNA-based vaccine, with reported IRR are 1.22, 1.21, 1.04, 1.13 and 1.08. The same design also had been used to evaluate the risk of those acute events after a SARS-CoV-2 infection, with IRR reported from other research are 63.52 (Venous thromboembolism) and 42.26 (Arterial thromboembolism) at the exposure date (8), 12.01 (thromboembolism), and 8.44 (acute myocardial infarction) and 6.18 (ischaemic stroke) in the first week after exposure (11), (14), and result obtained from our database is 23.46 in 28 days after a positive PCR test. However, as the current evidence are based on datasets sampled from hospitalized individuals, who were more likely to take the COVID-19 test, there is an underrepresentation and lack of generalizability to the broader population (20). In other words, SCCS suffers from misclassification issues, since subjects without any acute event can be misclassified into the noninfection category. Therefore, the estimated relative risk of thrombosis after a SARS-CoV-2 infection obtained by SCCS is indeed inflated.

The objective of this study is to evaluate the overall risks and benefits associated with SARS-CoV-2 vaccination on the occurrence of thromboembolism events. To do so, we first quantified the risk of acute thromboembolism after a mRNA-based vaccine using the SCCS method. Secondly, we evaluated the association between the acute thromboembolism events with a SARS-CoV-2 infection using a case-control study, which was expected to tackle the bias due to misclassification issue. Finally, we examined the mechanism underlined in the relationship between the occurrence of thrombosis and COVID-19 vaccination, exploring the impact of vaccination on COVID-19 infection, thereby indirectly influences the probability of experiencing an adverse event in vaccinated group.

## 2. Method

### 2.1 Data

We used Electronic Health Record (EHR) of Corewell Health East healthcare system in this study. We included patients who were registered with a primary care physician within 18 months prior to Jan 1^st^ 2021, and had any outcome of interest within the observation period (Dec 1st 2020 – August 31st 2022). We identified acute thrombosis events by a hospital admission or death associated with venous thromboembolism or arterial thromboembolism, see Table 4 in Appendix for a list of ICD-10 (International Classification of Diseases version 10) diagnosis codes. We further excluded individuals who were under 18 on Jan 1^st^ 2021, had incomplete covariate data, or had a hospital admission for thrombosis within 365 days before the first event in the observation period.

Our primary interest is to study the mRNA-based vaccines effect on the occurrence of thrombosis using the SCCS design. We included patients who had at least the first vaccine dose of mRNA-1273 (Moderna) or BNT162b2 (Pfizer) and experienced any outcome of interest. We further excluded patients who had an infection 90 days prior to any dose of vaccination. We used the EHR data system to identify vaccine exposure, which includes the date of vaccination, vaccine type and dose order.

The same SCCS was used to study the risk of thromboembolism associated with a SARS-Cov-2 infection, in which we included patients who had at least one positive PCR test and a thrombosis event during the observational period. However, as a result of misclassification, the SCCS derived an inflated IRR 23.46. Therefore, we proposed a case-control design, in which we included patients who either had a thrombosis event or hospitalized injury in the observational period, (see Table 4 in Appendix for a list of ICD-10 codes), and a SARS-CoV-2 (PCR or antigen) test or within 28 days period prior to the hospital admission date. We determined SARS-CoV-2 infection status based on the test results during these 28 days period. If an individual only had negative PCR or antigen test results, this subject was classified into noninfection. Moreover, we identified disease status by the earliest event, in which patients having thrombosis is classified into case group. Additionally, if an individual had an admission for both thrombosis and injury, then this subject would be considered as having thrombosis. In this way, each of individual were classified into one disease status (case-control).

### 2.2 Statistical Method

We used self-controlled case series (SCCS) design to examinate the association of acute thromboembolism and the first two doses of mRNA-based COVID-19 vaccines (mRNA-1273 or BNT162b2) from December 1^st^ 2020 to August 31^st^ 2022. The SCCS method compares the incidence of thromboembolism after and before vaccination. In this method, subjects are their own control and comparisons are made within subjects, therefore, all time invariant confounding is removed. We included subjects who had a thrombosis event and had at least the first vaccine dose in the study period. The control period (i.e., unexposed period) is defined from December 1^st^ 2020 to 28 days prior to the first dose of vaccination. We excluded the 28 days prior vaccination to avoid bias due to contra-indications (21). Two separate risk periods for the first and second dose were defined from the vaccination date to 28 days, the date of infection or death, whichever occurred first. See Figure 1 in the Appendix. We also excluded subjects who had COVID-19 infection within 90 days before the first vaccine dose to remove the confounding effect of infection on the outcome. To ensure independence between repeatedly occurred outcomes, we considered outcome events occurred at least one year after the previous event. We used the conditional Poisson regression including an offset for the length of risk period to obtain the incidence rate ratio (IRR) of thrombosis after and before COVID-19 vaccination.

**Figure 1.**
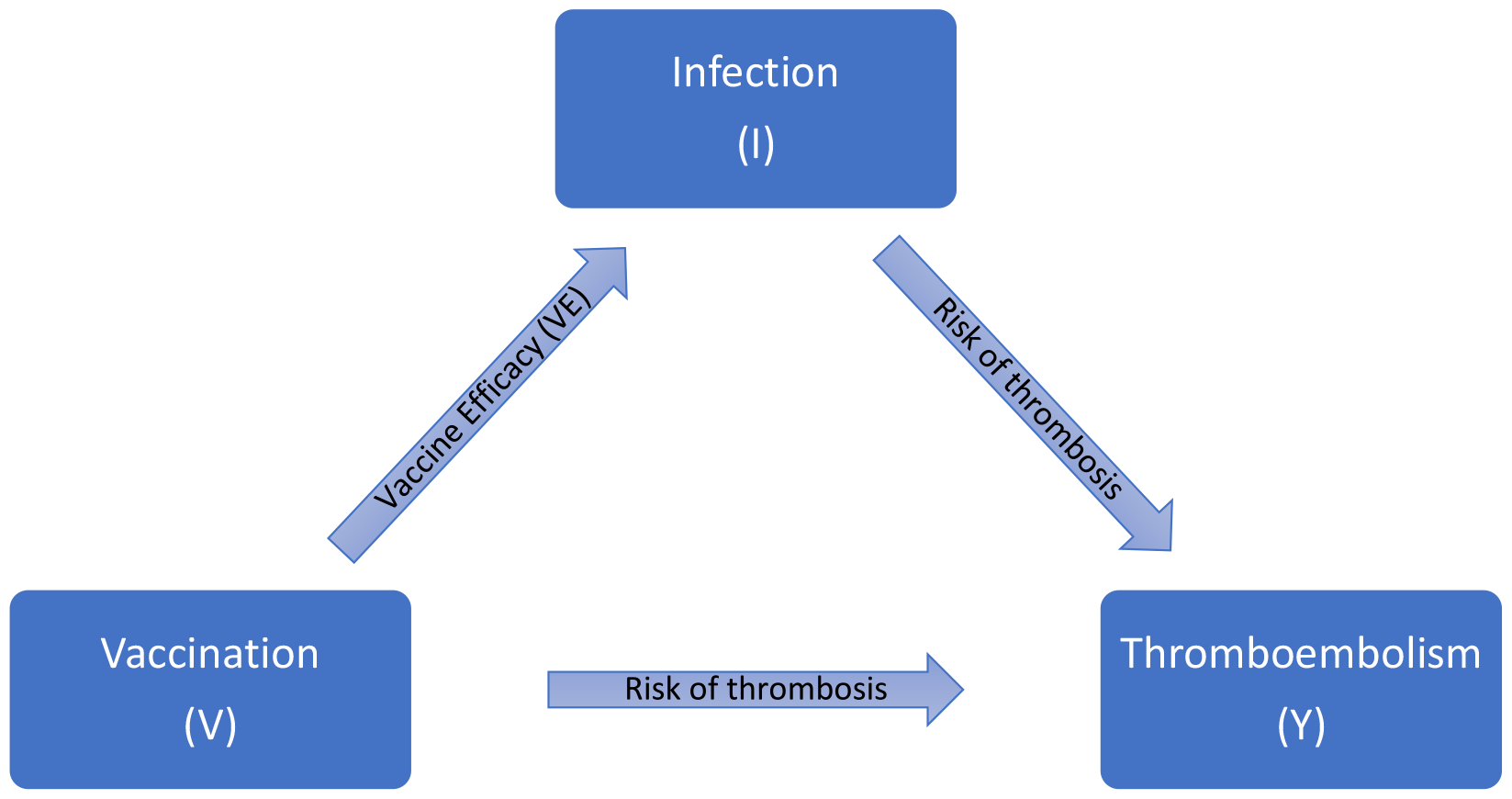
Direct and indirect association between mRNA vaccine and thromboembolism.

It is challenging to study the association between the adverse events and the COVID-19 infection due to inaccurate infection data in patients who did not have the thrombosis event. Patients visited emergency department or before admitted to hospital, were likely to take a COVID-19 test (antigen or PCR) especially during early pandemic. Therefore, observing event of thromboembolism is more likely to be associated with a COVID-19 infection because subjects with adverse events is more likely to take the COVID-19 test compared to subjects without any adverse event, even if there is no association between the acute events and an infection. The under-estimated infection rate in controls group leading to the overestimation of the risk of thromboembolism after a COVID-19 infection [estimated IRR is 23.46 showing an inflation]. This fact is evidenced by the analysis of the hospitalized injury as the outcome, which has the same misclassification issues, but is not associated with COVID-19 infection, we found IRR is 10.35.

The main idea is to address the misclassification problem in the control subjects (i.e., subjects without thromboembolism). We proposed a simple and efficient method to quantify the association while dealing with the misclassification issue. Early in the pandemic, everyone who visited ED or before admitted to hospital were tested for COVID-19. Therefore, we assumed subjects who had a thrombosis event had correct infection data (i.e., test has 100% accuracy), and the misclassification only occurred in subjects without thrombosis (control subjects). The idea is to select a subset of control group whose infection data is certain. To this end, we selected event of hospitalization related to injury, as injury is not associated with COVID-19 infection and hospitalized event had correct infection data, we selected it as the control group, and obtained the odd ratio measuring ratio of the odd of thrombosis and inpatient injury after and before a positive test.

In the case-control design, we compared the odds of infection (exposed) vs uninfection (unexposed) in the cases (thromboembolism) vs. controls (injury). If an individual has a thrombosis/injury (case/control) event, we considered this subject is infected by SARS-CoV-2 (exposed) if they had a positive PCR or antigen test within 28 days before the event. Moreover, to further avoid the possible systematic error caused by unreported infection, only patients who had all negative PCR test result in the period of 28 days before the occurrence of event is considered as uninfected (unexposed). Logistic regression models were adjusted for age, race, gender, Charlson comorbidity index (CCI), and prior vaccination status (Yes/No). The CCI was obtained using the R package comorbidity and categorized into 4 categories, ’0’, ’1-2’, ’3-4’, and ’≥ 5’ (22), (23). We reported Odd Ratios (OR), 95% confidence interval (CI).

Statistical analyses were performed in R 4.3.0. OR and IRR estimates were presented with 95% CIs and p-value. We used de-identified EHR data, the use of which was approved by the Institutional Review Board (IRB) of Corewell Health.

## 3. Result

### 3.1 Study population

During the study period, there were 448524 COVID-19 vaccinated subjects in Corewell Health East data base, among which 170960 (38.12%) had the first two doses of mRNA-1273 and 277564 (61.88%) took BNT162b2. Overall, the number of patients who were fully vaccinated was 427458 (95.30%), and there were 21066 (4.70%) patients having only one dose. The median age is 57 (with interquartile range [IQR]: 42 – 69), and 60.26% patients were female and 39.74% are male.

Among those vaccinated, there were 12415 patients, who had at least one event and had the first dose of either mRNA-1273 or BNT162b2 vaccine. Moreover, these patients did not have any SARS-CoV-2 infection within 90 days before vaccination or between vaccination date and date of event occurrence, and they also had a clean one-year medical history, so that the first hospital admission relating to thrombosis in observational period is the newest within one year. Demographic of patients in the study of vaccination is presented in Table 2 in Appendix. We identified 16722 events in the observational period, among which there were only 2139 thrombosis occurred in the control period, 527 and 575 events in the risk periods after the first and second dose of vaccination, see Table 1 in Appendix. In conclusion, there were only 2798 patients and total of 3241 events were considered in our conditional Poisson regression.

During observational period, there were 26052 patients receiving a positive PCR/antigen test result. The median age is 46 (with IQR: 28-62), and 58.63% are female and 41.37% are male.

There were 20249 patients who got a hospital admission relating to either thromboembolism or injury during the observational period. These patients had at least one COVID-19 test recorded in 28 days prior to the hospital admission. There were 43.04% patients in control group (injury) and the rest of 56.96% are in the case group (thromboembolism). Demographic of patients in the study of infection is presented in Table 3 in Appendix.

### 3.2 The association between thromboembolism and exposures

#### Thromboembolism and mRNA-vaccination

We found an increased risk of thrombosis after the first dose (IRR is 1.16 with 95% CI: [1.04, 1.29]), and after the second dose (IRR is 1.19 and 95% CI: [1.07,1.32]) of the mRNA-base vaccination.

#### Thromboembolism and SARS-Cov-2 exposure

We found a significant interaction between SARS-Cov-2 exposure and prior vaccination status. The OR of experiencing a thromboembolism event after a SARS-Cov-2 infection depends on whether if the patient had any dose of vaccine before the infection. The reported OR among unvaccinated and vaccinated group are 1.77 (CI: [1.48, 2.1]) and 1.34 (CI: [1.09, 1.66]) respectively. We observed increased risks of thrombosis after a SARS-CoV-2 infection in both groups, however vaccination seems to have some protection against adverse events as the OR estimated in this group is lower, compared to the group of unvaccinated.

### 3.3 Estimate the overall effect of vaccination on thromboembolism while considering infection

We extended our investigation to evaluate the overall influence of COVID-19 vaccination on the occurrence of acute thromboembolism events. COVID-19 vaccines dramatically reduce the SARS-Cov-2 infection rate (24), (25), (26), hence potentially indirectly decreases the likelihood of experiencing an acute thromboembolism event among the vaccinated group. Figure 1 below illustrates the direct and indirect effect of the COVID-19 vaccination on thromboembolism while considering the vaccine efficacy (VE). As presented in the diagram, the association between thromboembolism and COVID-19 vaccination is described by two paths, the direct association represents the direct increased risk of thromboembolism after any dose of vaccination, and the indirect association between thrombosis and vaccination is represented by the potential reduction in the risk of thromboembolism through the decreased risk of COVID-19 infection by vaccination. We estimated the overall influence of vaccination on the occurrence of thromboembolism by considering both direct and indirect paths.

For a vaccinated subject, the total risk is P(Y|V, Ī) + P(I|V) × P(Y|I, V), where P(Y|V, I) is the direct risk of thromboembolism after vaccination given that infection has not yet occurred, and P(I|V) × P(Y|I, V) is the indirect risk calculated by multiplying the risk of COVID-19 infection of a vaccinated subject, P(I|V), and the risk of thromboembolism given a COVID-19 infection in the vaccinated group, P(Y|I, V). Similarly, the overall risk of thromboembolism for an unvaccinated subject is as 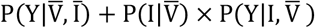. Hence the relative risk of thrombosis for a vaccinated subject compared to an unvaccinated subject is

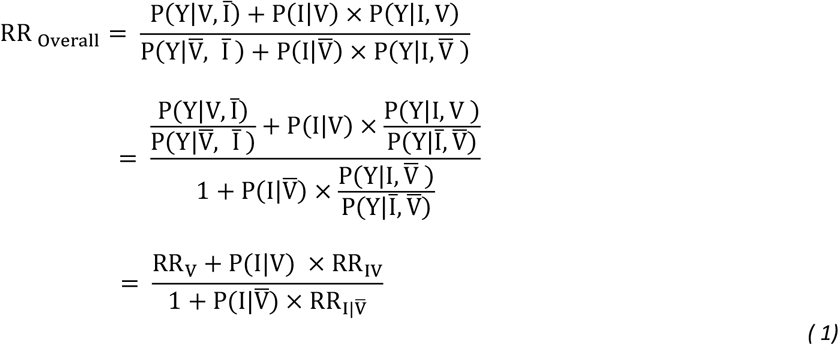

The terms RR_V_ is the relative risk of thromboembolism associated with COVID-19 vaccination, and 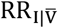 is the relative risk of thromboembolism after a COVID-19 infection in the group of unvaccinated. The term RR_IV_ is relative risk of thromboembolism between the group having both type of exposures, vaccination, and then infection, compared to unexposed group. Our analysis with the Corewell Health EHR data in the previous section gave an IRR of 1.19 as the measure of the association between thromboembolism and COVID-19 vaccination, therefore, RR_V_ = 1.19. We also obtained an odd ratio 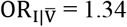 and OR_IV_ = 1.43 from the analysis using case-control design. We therefore set 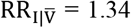 and RR_IV_ = 1.43 as the event of thromboembolism is rare (27). Hence, the above overall relative risk becomes

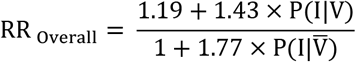

The overall relative risk of thromboembolism after a SARS-CoV-2 vaccination depends on the infection rate for both vaccinated and unvaccinated subjects. We defined vaccine efficacy 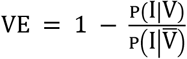, then with a given infection rate for an unvaccinated subject, 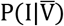, we derived risk of infection for a vaccinated subject as 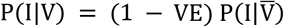, and obtained the overall risk, RR _Overall_, as a function of VE.

Figure 2 illustrates the overall relative risk of thromboembolism after a mRNA-vaccination as a function of VE. As VE increases from 0 to 1, the overall risk decreases. Moreover, infection rate, 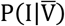, also affects the reduction of the risk of thromboembolism. During the period with higher infection rate, the benefit of vaccination is stronger. More specifically, with VE is .8, the overall risk of thromboembolism after vaccination is decreased by 10% during the period of infection rate .2, while it decreased only 3% during the period of infection rate .15.

**Figure 2.**
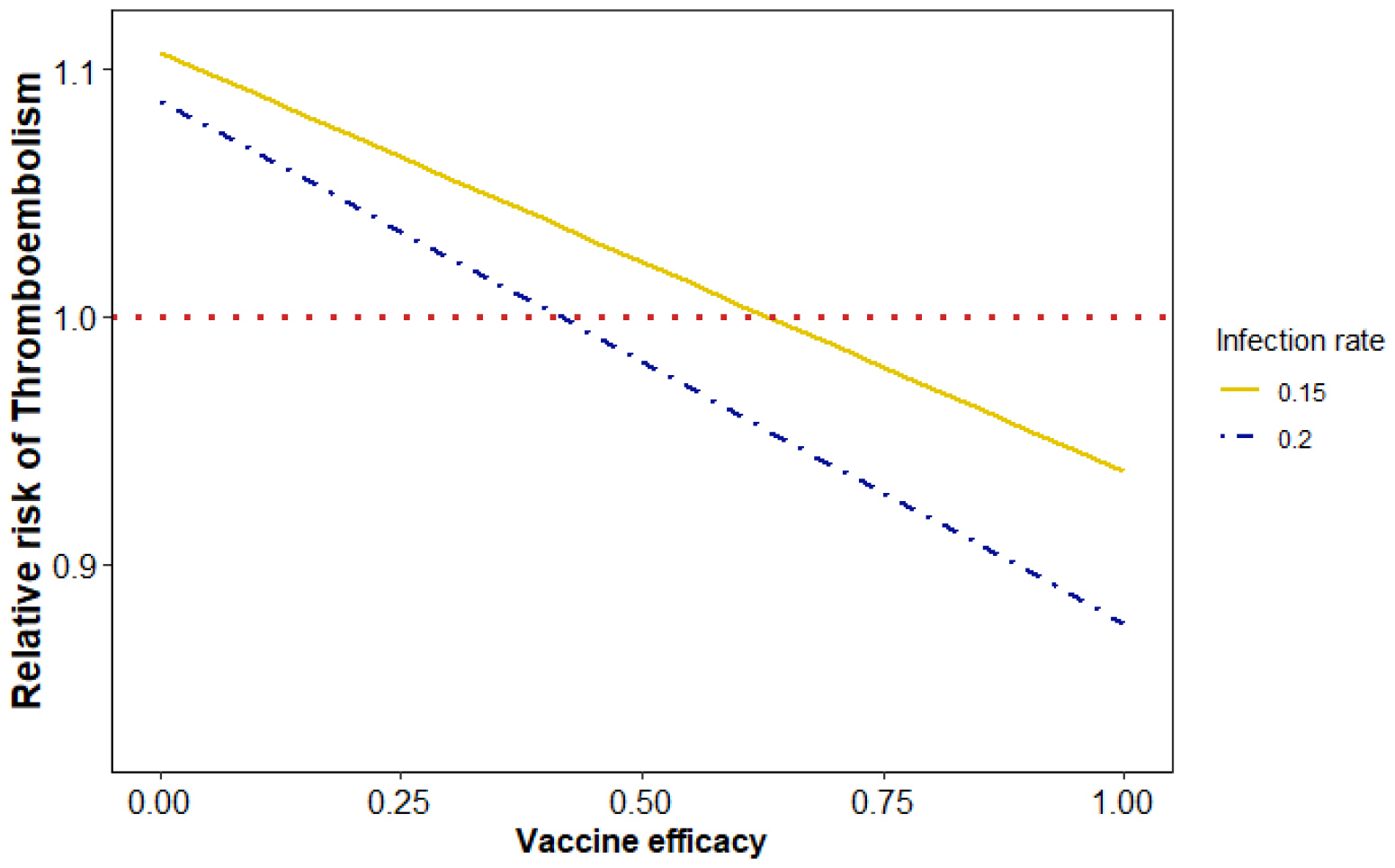
Overall risk of thromboembolism, comparison between vaccinated and unvaccinated groups. Infection rate .15 and .2 is the probability of SARS-Cov-2 infection before vaccination (27).

## 4. Discussion

We found that both COVID-19 vaccination and COVID-19 infection increase risk of thromboembolism. However, evidence implies that the probability of experiencing a thrombosis event after a SARS-Cov-2 infection is higher, compared to the exposure to a mRNA-base vaccination. Our analysis is in agreement with previous research, indicating that adverse event followed by a COVID-19 infection would be more severe, compared to vaccination (8), (12), (13), (14).

This analysis is subject to a few limitations. First, we considered the newest event within a one-year medical history, hence the correlation of events across exposure group is low while within an exposure period is still high. In the view that risk period is restricted for at most 28 days, and control period usually is longer, this strategy can possibly overestimate the relative risk ratio. Additionally, our case-control design also has control selection bias as inpatient injury is only a subset of the control cohort and it may not represent the cohort, which may produce a biased estimate of the odd ratio. Evidence for this bias is the non-significant odd ratio in the study of the effect of mRNA-vaccination using the same case-control design (OR after the first dose is 1.02, CI: [.95, 1.09]), and OR after the second dose is 1.0 with CI: [.94, 1.06]) while results from SCCS indicated a positive correlation with IRR (1.16 and 1.19 after the first and second dose). Therefore, the selection bias in case-control design implied an underestimation of OR and the true association would be more significant. Hence, the overall risk of thromboembolism followed by a COVID-19 vaccination expressed in Equation (1) should have a stronger decrease, and the reduction of probability of experiencing a thromboembolism event in vaccinated group should be more significant.

## 5. Conclusion

The analysis provides evidence of the increased risk of thromboembolism event within a short-term risk period after a SARS-Cov-2 vaccination. However, it becomes evident that the benefits outweigh the risks. Patients with prior vaccination have a lower risk of thromboembolism associated with a SARS-Cov-2 infection, compared to non-vaccinated patients. Moreover, the benefit of COVID-19 vaccination manifests through its role in preventing and controlling the spread of the SARS-CoV-2 virus, and further decreases the incidence of thrombosis caused by infection, hence generally derives a reduction of probability of having an acuate thrombosis event among vaccinated population.

## Data Availability

All data produced in the present study are available upon reasonable request to the authors.

## Appendix

**Figure 1.**
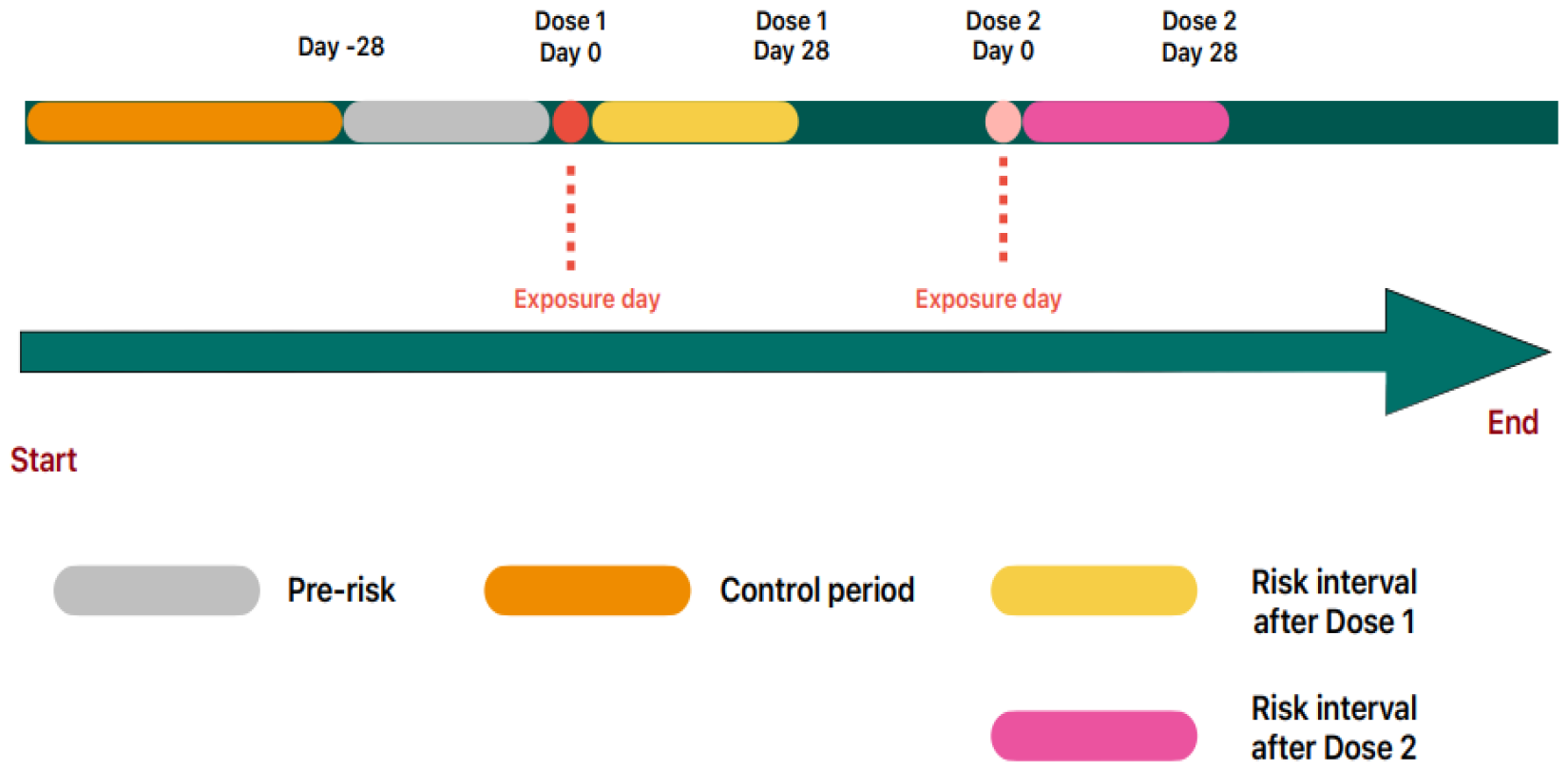
Self-controlled case series study design. Each patient is followed from the index date to study end date and censored if death of end of observational period.

**Table 1.** Distribution of thrombosis in the calendar time from 1 December 2020 to 31 August 2022.

| Group | Exposure Group | Number of events | IRR |
| --- | --- | --- | --- |
| mRNA Vaccine | Control period | 2139 | * |
|  | Not interest | 13064 | * |
|  | 28 days before the first dose (Pre-risk) | 417 | * |
|  | 0-28 days after dose 1 | 527 | 1.16 (1.04-1.29) |
|  | 0-28 days after dose 2 | 575 | 1.19 (1.07, 1.32) |
|  | Total events | 16722 |  |

**Figure 2.**
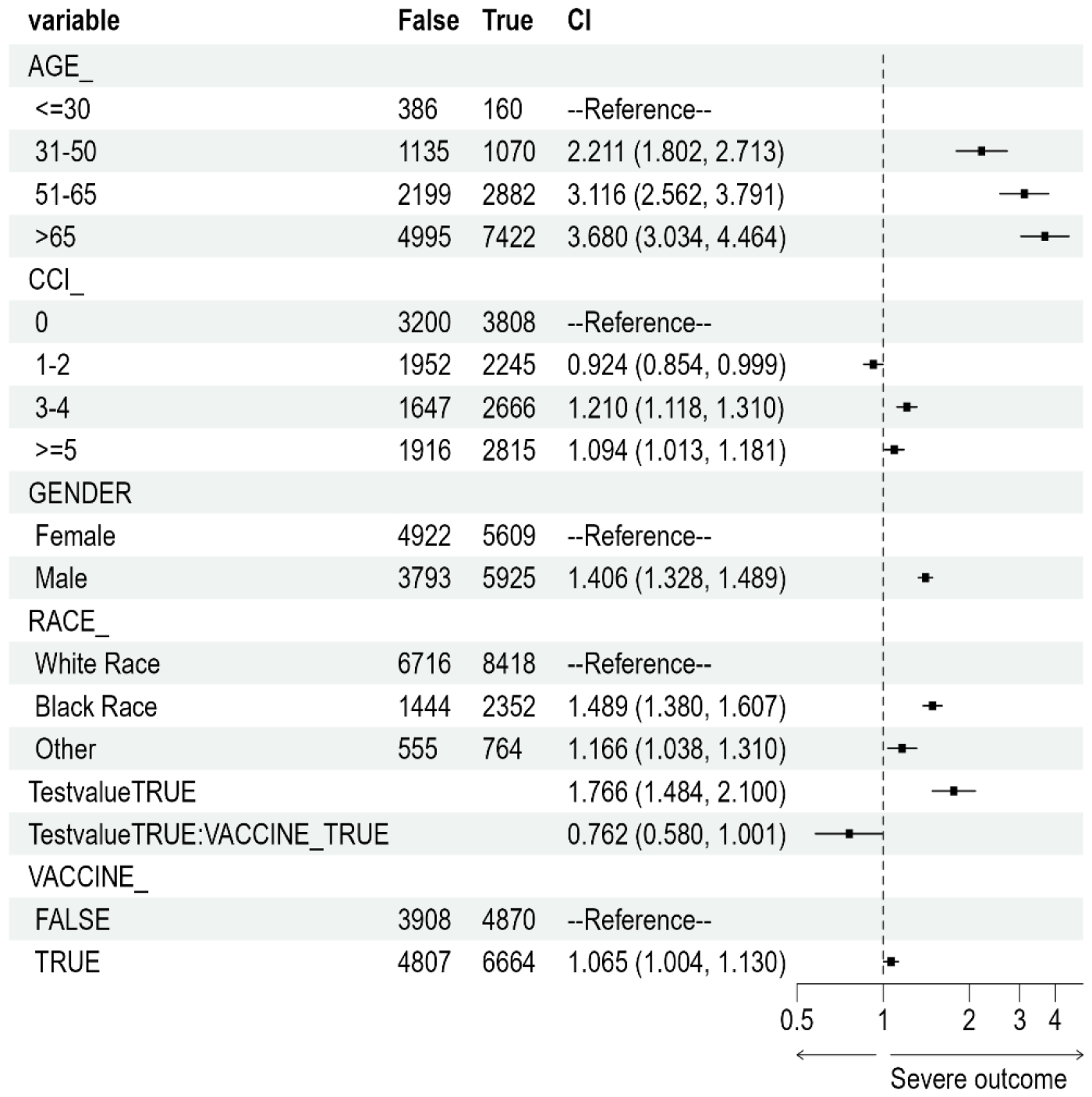
Forest plot of case-control design studying association of thrombosis and SARS-Cov-2 infection.

**Table 2.**
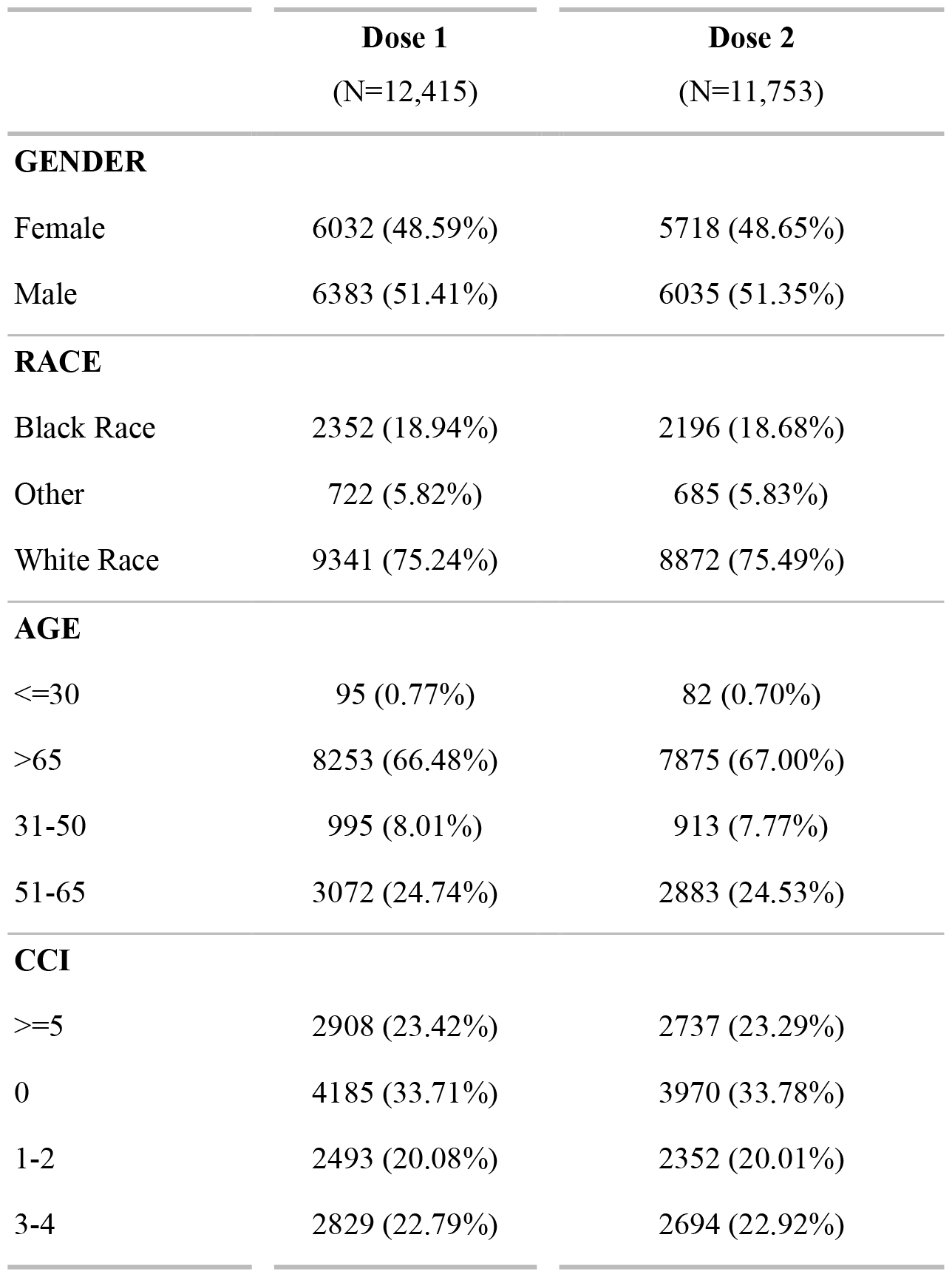
Demographic Characteristics of patients who experienced at least on thromboembolism event and had at least one dose of mRNA-vaccination from Dec 1^st^ 2020 to August 31^st^ 2022.

**Table 3.** Demographic Characteristics of patients who experienced at least on thromboembolism or inpatient injury event and had at least a PCR test from Dec 1^st^ 2020 to August 31^st^ 2022.

|  | **Injury**<br>(N=8,715) | **Thromboembolism**<br>(N=11,534) | **Overall**<br>(N=20,249) |
| --- | --- | --- | --- |
| **Gender** |  |  |  |
| Female | 4922 (56.48%) | 5609 (48.63%) | 10531 (52.01%) |
| Male | 3793 (43.52%) | 5925 (51.37%) | 9718 (47.99%) |
| **Race** |  |  |  |
| White Race | 6716 (77.06%) | 8418 (72.98%) | 15134 (74.74%) |
| Black Race | 1444 (16.57%) | 2352 (20.39%) | 3796 (18.75%) |
| Other | 555 (6.37%) | 764 (6.62%) | 1319 (6.51%) |
| **Age** |  |  |  |
| <=30 | 386 (4.43%) | 160 (1.39%) | 546 (2.70%) |
| 31-50 | 1135 (13.02%) | 1070 (9.28%) | 2205 (10.89%) |
| 51-65 | 2199 (25.23%) | 2882 (24.99%) | 5081 (25.09%) |
| >65 | 4995 (57.31%) | 7422 (64.35%) | 12417 (61.32%) |
| **CCI** |  |  |  |
| 0 | 3200 (36.72%) | 3808 (33.02%) | 7008 (34.61%) |
| 1-2 | 1952 (22.40%) | 2245 (19.46%) | 4197 (20.73%) |
| 3-4 | 1647 (18.90%) | 2666 (23.11%) | 4313 (21.30%) |
| >=5 | 1916 (21.99%) | 2815 (24.41%) | 4731 (23.36%) |
| **Prior Vaccination** |  |  |  |
| FALSE | 3908 (44.84%) | 4870 (42.22%) | 8778 (43.35%) |
| TRUE | 4807 (55.16%) | 6664 (57.78%) | 11471 (56.65%) |

**Table 4.** List of ICD10 codes.

| Outcome | ICD10 code | ICD10 code and description |
| --- | --- | --- |
| **Venous thromboembolism** | I26 | Pulmonary embolism |
|  | I260 | Pulmonary embolism with mention of acute cor pulmonale |
|  | I269 | Pulmonary embolism without mention of acute cor pulmonale |
|  | I81 | Portal vein thrombosis |
| **Venous thromboembolism**<br>**Venous thromboembolism**<br>**Arterial thrombosis** | I81X | Portal vein thrombosis |
|  | I82 | Other venous embolism and thrombosis |
|  | I820 | Budd-Chiari syndrome |
|  | I822 | Embolism and thrombosis of vena cava |
|  | I823 | Embolism and thrombosis of renal vein |
|  | I828 | Embolism and thrombosis of other specified veins |
|  | I829 | Embolism and thrombosis of unspecified vein |
|  | I74 | Arterial embolism and thrombosis |
|  | I740 | Embolism and thrombosis of abdominal aorta |
|  | I741 | Embolism and thrombosis of other and unspecified parts of aorta |
| **Venous thromboembolism** | I742 | Embolism and thrombosis of arteries of upper extremities |
| **Arterial thrombosis**<br>**Cerebral venous sinus thrombosis** | I743 | Embolism and thrombosis of arteries of lower extremities |
|  | I744 | Embolism and thrombosis of arteries of extremities, unspecified |
|  | I745 | Embolism and thrombosis of iliac artery |
|  | I748 | Embolism and thrombosis of other arteries |
|  | I749 | Embolism and thrombosis of unspecified artery |
|  | O225 | Cerebral venous thrombosis in pregnancy |
|  | O873 | Cerebral venous thrombosis in the puerperium |
|  | I636 | Cerebral infarction due to cerebral venous thrombosis |
|  | I676 | Non-pyogenic thrombosis of intracranial venous system |
| **Ischaemic stroke** | G45 | Transient cerebral ischaemic attacks and related syndromes |
|  | G450 | Vertebrobasilar artery syndrome |
|  | G451 | Carotid artery syndrome (hemispheric) |
|  | G452 | Multiple and bilateral precerebral artery syndromes |
| **Ischaemic stroke**<br>**Myocardial infarction** | G453 | Amaurosis fugax |
|  | G454 | Transient global amnesia |
|  | G458 | Other transient cerebral ischaemic attacks and related syndromes |
|  | G459 | Transient cerebral ischaemic attack, unspecified |
|  | I63 | Cerebral infarction |
|  | I630 | Cerebral infarction due to thrombosis of precerebral arteries |
|  | I631 | Cerebral infarction due to embolism of precerebral arteries |
|  | I632 | Cerebral infarction due to unspecified occlusion or stenosis of precerebral arteries |
|  | I633 | Cerebral infarction due to thrombosis of cerebral arteries |
|  | I634 | Cerebral infarction due to embolism of cerebral arteries |
|  | I635 | Cerebral infarction due to unspecified occlusion or stenosis of cerebral arteries |
|  | I636 | Cerebral infarction due to cerebral venous thrombosis, non-pyogenic |
|  | I638 | Other cerebral infarction |
|  | I639 | Cerebral infarction, unspecified |
|  | I64 | Stroke, not specified as haemorrhage or infarction |
|  | I64X | Stroke, not specified as haemorrhage or infarction |
|  | I21 | Acute myocardial infarction |
|  | I210 | Acute transmural myocardial infarction of anterior wall |
|  | I211 | Acute transmural myocardial infarction of inferior wall |
|  | I212 | Acute transmural myocardial infarction of other sites |
| **Myocardial infarction** | I213 | Acute transmural myocardial infarction of unspecified site |
|  | I214 | Acute subendocardial myocardial infarction |
|  | I219 | Acute myocardial infarction, unspecified |
|  | I22 | Subsequent myocardial infarction |
|  | I220 | Subsequent myocardial infarction of anterior wall |
|  | I221 | Subsequent myocardial infarction of inferior wall |
|  | I228 | Subsequent myocardial infarction of other sites |
|  | I229 | Subsequent myocardial infarction of unspecified site |
|  | I23 | Certain current complications following acute myocardial infarction |
|  | I230 | Haemo-pericardium as current complication following acute myocardial infarction |
|  | I231 | Atrial septal defect as current complication following acute myocardial infarction |
|  | I232 | Ventricular septal defect as current complication following acute myocardial infarction |
|  | I233 | Rupture of cardiac wall without haemopericardium as current complication following acute myocardial |
|  | I234 | Rupture of chordae tendineae as current complication following acute myocardial infarction |
|  | I235 | Rupture of papillary muscle as current complication following acute myocardial infarction |
|  | I236 | Thrombosis of atrium, auricular appendage, and ventricle as current complications following acute myocardial |
|  | I238 | Other current complications following acute myocardial infarction |
|  | I252 | Old myocardial infarction |
| **Injuries** | S00-S09 | Injuries to the head |
|  | S10-S19 | Injuries to the neck |
|  | S20-S29 | Injuries to the thorax |
|  | S30-S39 | Injuries to the abdomen, lower back, lumbar spine, pelvis and external genitals |
|  | S40-S49 | Injuries to the shoulder and upper arm |
|  | S50-S59 | Injuries to the elbow and forearm |
|  | S60-S69 | Injuries to the wrist, hand, and fingers |
|  | S70-S79 | Injuries to the hip and thigh |
|  | S80-S89 | Injuries to the knee and lower leg |
|  | S90-S99 | Injuries to the ankle and foot |
|  | T07-T35 | Injures and others consequences of external causes |
|  | T51-T88 |  |

